# Characterizing the informativeness of pathogen genome sequence datasets about transmission between population groups

**DOI:** 10.1101/2025.08.07.25333239

**Authors:** Cécile Tran-Kiem, Amanda C. Perofsky, Justin Lessler, Trevor Bedford

## Abstract

Pathogen genome analysis helps characterize transmission between population groups. The information carried by pathogen sequences comes from the accumulation of mutations within their genomes; thus, the pace at which mutations accumulate should determine the granularity of transmission processes that pathogen sequences can characterize. Here, we investigate how the complex interplay between mutation, transmission, population mixing and sampling impacts study power. First, we develop a conceptual probabilistic framework to quantify the ability of pairs of sequences in capturing between-group transmission history. This allows us to comprehensively explore the space of possible phylogeographic analyses by explicitly considering the pace at which mutations accumulate and the pace at which between-group transmission events occur. Using this framework, we identify a pathogen-intrinsic limit in the mixing scale at which their sequence data remains informative, with faster mutating pathogens enabling finer spatial characterization. Secondly, we perform a simulation study exploring a range of assumptions regarding sequencing intensity. Sample size further imposes a limit on the characterization of between-group transmission processes. This work highlights inherent horizons of resolvability for population mixing processes that depend on the interaction between evolution, transmission, mixing and sampling. Such considerations are important for the design of pathogen genomic studies.

## Introduction

Pathogen sequencing is an invaluable tool for studying disease transmission patterns^1^. Analyzing pathogen genomes alongside metadata describing characteristics of infected hosts from which pathogens were isolated has helped characterize the transmission of pathogens between population groups of varying sizes. For example, hemagglutinin phylogenies have shed light on the inter-continental spread of seasonal influenza viruses^1^, and identical sequences occurrence patterns have illuminated SARS-CoV-2 transmission between age groups^2^.

It is generally acknowledged that the fundamental power of genomic epidemiological studies arises from the fast pace at which genetic variation is generated within pathogen genomes^3^, relative to the pace at which pathogens transmit between hosts. When transmission and mutation events occur over similar timescales, pathogen genomes isolated from infected hosts indeed contain information about underlying epidemiological processes^4^. Thus, analysing such genomic data can help reconstruct transmission chains^5^ or characterize population-level patterns of pathogen spread^1,2^. Prior work has explored the ability of pathogen genomes to reconstruct transmission chains^3,5^, with overall resolution depending on evolutionary rate, generation time, transmission intensity and sampling effort. However, we still lack clear methods to evaluate both the power and limits of pathogen genome sequences in quantifying transmission at the population level (phylogeographic inference). Making such capabilities and limits explicit is important to guide study design, set realistic expectations about genomic epidemiology’s role for epidemic response and ensure the efficient use of sequencing resources.

To study such population processes, we expect the relative timescale at which mutation^6^ and between-group transmission events occur to be critical^7^. This is because we anticipate pathogen genomes to be informative about a process only up to the rate at which novel genomic variation is observed^6,7^. If mutations accumulate at a much slower pace than between-group transmission events occur (high between-group transmission / low mutation rate in Figure 1), genome sequences are insufficient to infer between-group transmission patterns, as highlighted by the presence of large well-mixed polytomies in the phylogeny. Analyzing sequences from a faster mutating pathogen might enable characterization of such a between-group transmission process: although population mixing occurs rapidly, genome sequences aren’t sufficiently divergent to capture between-group transmission patterns (high between-group transmission / high mutation rate). Though insufficient to characterize rapid mixing processes, sequences from slow mutating pathogens still have the potential to decipher slow between-group transmission processes (low between-group transmission / low mutation rate).

**Figure 1:**
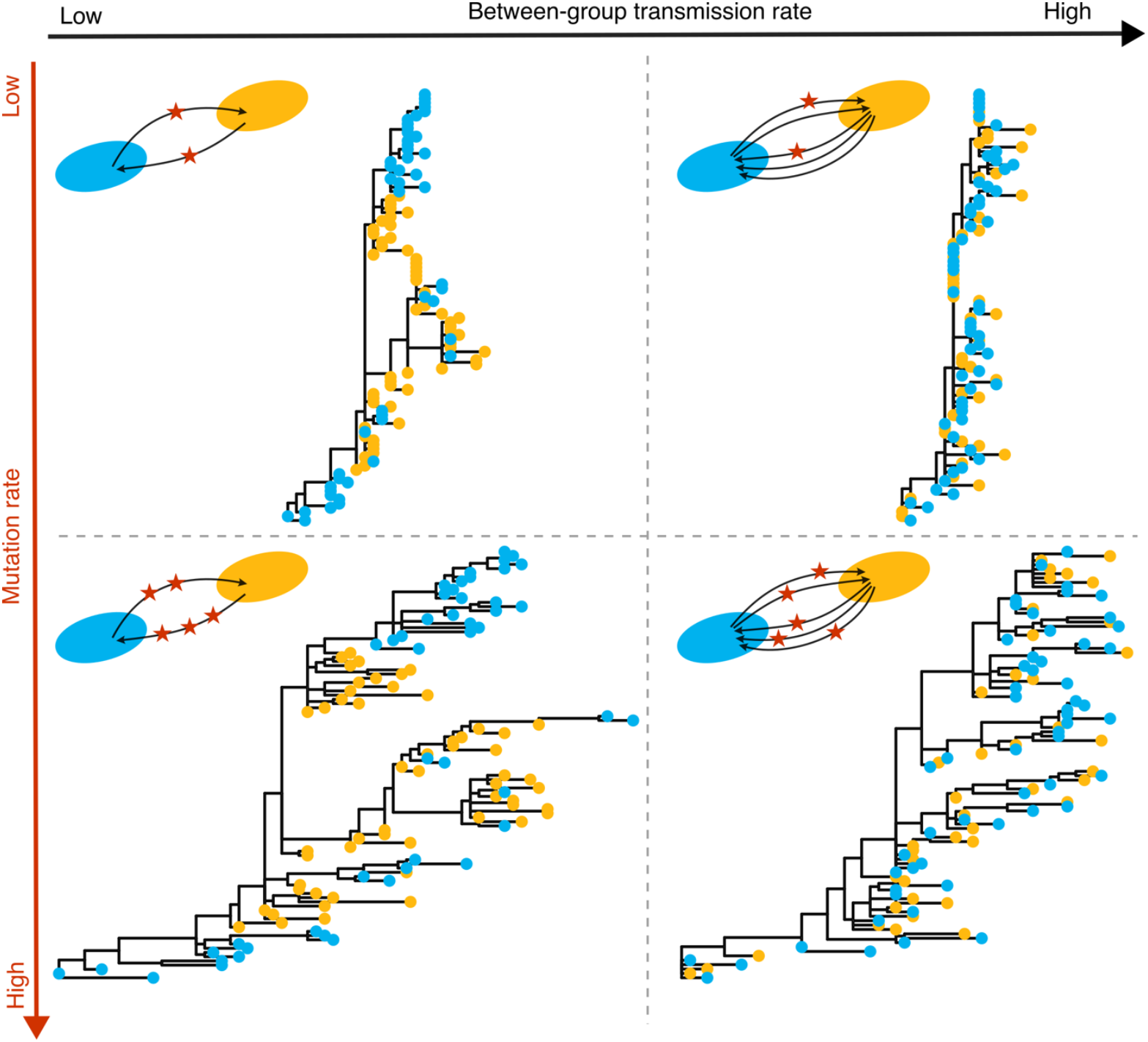
The ability of pathogen sequence data to characterize between-group transmission is impacted by the pace at which both between-group transmission and mutation events occur. To generate these figures, we simulate sequence data under an SEIR epidemic spreading between two groups of size 1000 (yellow and blue), assuming a genome length of 3,000 bp. We assume a basic reproduction number of 1.5, that the mean time spent in the exposed compartment is 3 days, and that the mean time spent in the infectious compartment is 3 days. 10% of infections is sequenced. We simulate the evolutionary process for a low mutation rate scenario (2·10^-5^ mutations/bp/day) and a high mutation rate scenario (8·10^-5^ mutations/bpday). For the mixing process, infected individuals have a 98% probability of transmitting to someone within their group in the low between-group transmission scenario and a 50% probability in the high between-group transmission scenario. For each scenario, we include on the top left a toy figure to illustrate the frequency of between-group transmission events (number of arrows) and mutation events (number of stars).

Here, we study how the interaction between sampling, transmission and evolutionary processes impacts our ability to characterize transmission between population groups from sequence datasets, focusing on estimating between-group transmission rates. We focus on sequence data analysis at the consensus level. Such datasets can be analysed through phylogeny-based methods. However, establishing how these factors affect the power of tree-based methods (e.g. by influencing coalescent patterns) is delicate. Recent work has illustrated that genetic distance-based approaches can characterize transmission at the population level^2^. Such approaches provide a more straightforward angle to characterize how these many factors influence the information contained by sequence datasets. Here, we develop a conceptual framework describing the ability of characterizing pathogen spread between population groups from the analysis of pairs of genetically proximal sequences. We apply this framework to a range of pathogens, characterized by distinct evolutionary characteristics and natural history parameters, and mixing processes (between age groups and various geographic scales). From this, we identify fundamental limits in the ability of pathogen genome sequencing to capture transmission dynamics at the group level. Finally, we conduct a simulation study to characterize how sampling additionally impacts the signal contained by clusters of proximal sequences.

## Methods

### Problem framing

We quantify the extent to which pathogen genome sequencing is informative about between-group transmission processes. We define genomic *linkage criteria* between groups and evaluate how well they capture (true) transmission links. Specifically, we assume that two groups are genomically linked through an observed sequence pair if the genetic distance between the elements of this sequence pair lies below a defined threshold Δ. We want to determine what is the sensitivity *η*_Δ_, specificity *χ*_Δ_, positive predicted value *ϕ*_Δ_ and accuracy *A*_Δ_ of this group linkage criterion.

### Probabilistic framework

#### Notation

Let *J* denote the number of between-group transmission events separating two infected individuals. We assume that they occur under a Poisson process of rate *λ*, where *λ* is the between-group transmission rate. In phylogeographic studies focusing on geographical units, *λ* is often referred to as the migration rate. By modelling transmission in a well-mixed population, where between-group transmission events occur independently and at a constant rate, we don’t account for potential network structure (which can be important in outbreak settings such as households, schools or workplaces). This assumption should be most relevant when studying population-level transmission, where the within-group transmission probability can be treated as constant along a transmission chain, and is in line with standard tree-based approaches (discrete trait analysis, structured coalescent and birth–death models), that also don’t account for any network structure.

Let *M* denote the number of mutations between the infecting pathogens of these two infected individuals. We assume that mutation events occur under a Poisson process of rate *μ*, where *μ* is the per genome mutation rate. Let *G* be a random variable denoting the number of generations separating these two individuals, which we define as the number of transmission events separating these individuals along a transmission chain. We assume that the generation time follows a Gamma distribution of shape *α* and scale *β*.

#### Distribution of the number of mutations conditional on the number of generations

Under these assumptions, the number of mutations *M* conditional on the number of generations *G* = *g* follows a negative binomial distribution of shape *r*_*M*|*g*_ and probability *p*_*M*| *g*_ parameters^2^:

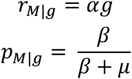

The full derivation is available in Supplementary Information.

#### Distribution of the number of between-group transmission events conditional on the number

*of generations*. Similarly, the number of between-group transmission events *J* conditional on the number of generations *G* = *g* follows a negative binomial distribution of parameters:

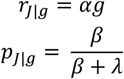

The full derivation is available in Supplementary Information.

#### Distribution of the number of between-group transmission events conditional on the number of mutations

In practice, we don’t observe the number of generations separating two infected individuals and are instead interested in the distribution of the number of between-group transmission events conditional on the number of mutations. Let *h*(*k*; *r, p*) denote the probability mass function evaluated in *k* of a negative binomial distribution of parameters *r* and *p*. We introduce *π*_*g*_ the probability for two sequenced individuals of being *g* generations apart. By integrating over the possible number of generations separating two infections, we show that:

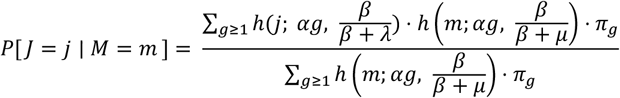

The full derivation is available in Supplementary Information. We assume each infection can only be sampled once, which means we don’t have any pair of samples in our dataset corresponding to *g* = 0 (0 transmission generation).

The distribution *π*_*g*_ of the number of generations between infected individuals is impacted by several factors, including the epidemic dynamics and the sampling scheme^8^. Wohl et al. used a simulation-based approach to approximate this probability distribution across a range of epidemiological scenarios, characterised by their reproduction number^8^. Their empirical estimates were obtained by simulating a branching process for *d* = ln(1000) /ln (*R*) generations, (where *R* is the reproduction number). This corresponds to the number of generations required to reach an expected epidemic size of 1000. From this, they derive the empirical distribution of the number of generations separating two infections (which we denote 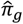). By design, the maximum number of generations separating two infected individuals in their simulations is therefore *g*_*max*_ = 2*d* which depends on the reproduction number.

We reuse the empirical probability distribution 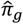 they estimated to fully approximate the probability *P*[ *J* = *j* ∣ *M* = *m*] as:

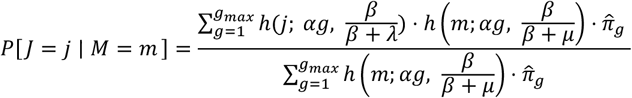

The probability of two infected individuals being separated by *j* between-group transmission events and *m* mutations thus follows:

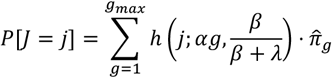

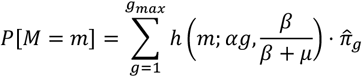

### Confusion matrix approach

#### Definition

To quantify the ability of a genetic linkage criterion to characterize transmission between population groups, we use a confusion matrix approach. We classify sequence pairs depending on their ability to accurately capture between-group transmission history from their genetic sequences. Figure 2 illustrates how we define the true between-group transmission history between two sequenced individuals and the inferred between-group transmission history from sequence data. For example, on the top left, the sequences of our two sampled infections define a linked pair (the number of mutations separating their genomes is below a predefined threshold). The true transmission history between these samples is *red* → *blue*. From our linked pairs, we infer that transmission occurred between these two groups (*red* ↔ *blue*). The inferred transmission history therefore accurately captures the true underlying history, corresponding to a True Positive (TP). A False Positive (FP) corresponds to a mismatch between the inferred between-group transmission history and the actual one. Likewise, if another between-group transmission event occurred between the pair members and the pair is not linked, that is a True Negative (TN), and if no other transmission event to another group occurred between the two sampled individual and the pair is not linked, that is a False Negative (FN). Our probabilistic framework enables to define the confusion matrix coefficients (Table 1). By definition, this linkage criterion doesn’t account for transmission direction and we focus on whether a sequence pair accurately represents the underlying between-group transmission history, regardless of directionality. Some pairs characterized by *J* > 1 may coincide with one segment of the full between-group transmission path (e.g. transmission *red* → *blue* → *red* → *blue* between two sequences, so that *red* ↔ *blue* captures a subset of the transmission history). We deliberately classify these pairs as TN or FP (like other pairs with *J* > 1) as they don’t capture the entire between-group transmission history. This formulation provides a conservative assessment of our linkage criterion’s performance.

**Table 1:**
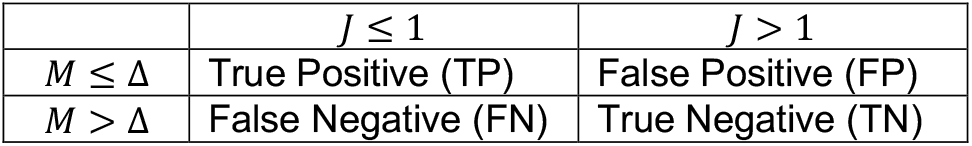
Confusion matrix coefficient for a linkage criterion based on a genetic distance threshold Δ.

**Figure 2:**
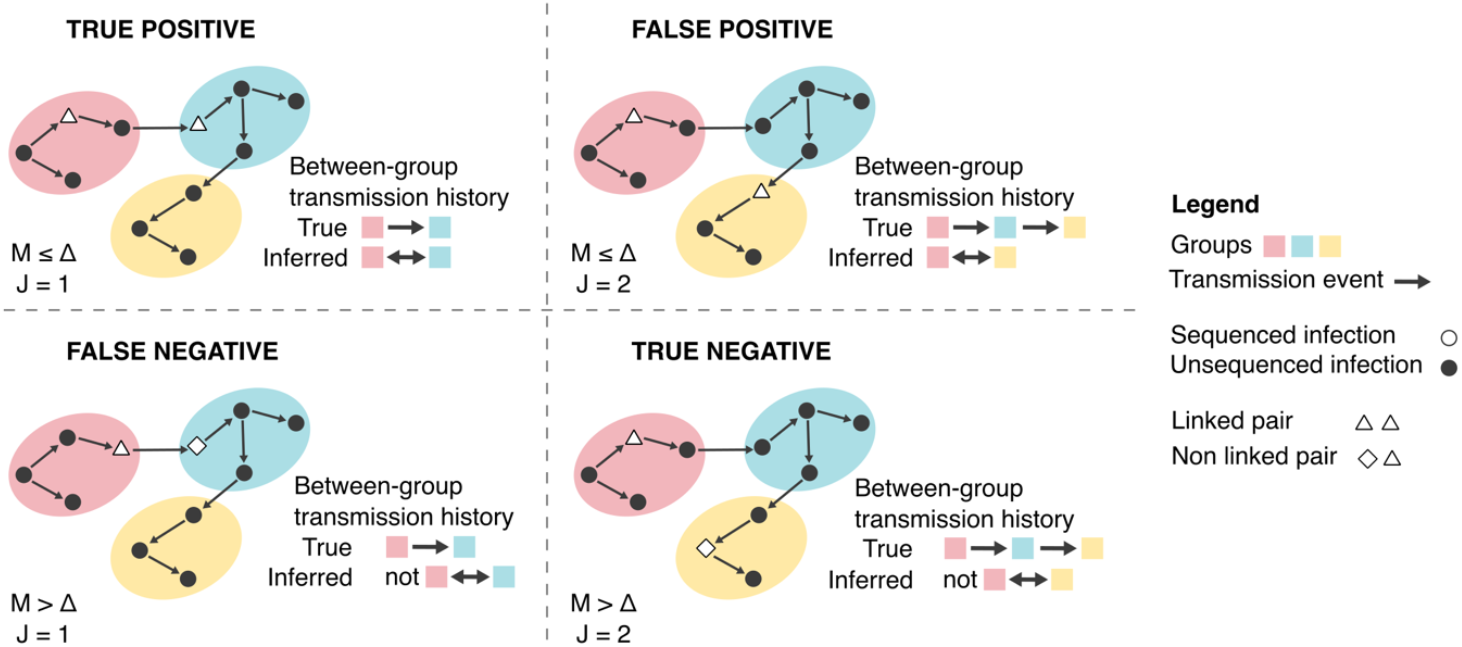
Illustration of the confusion matrix used to describe the ability of a genetic linkage criterion to capture the pathogen’s between-group transmission history. These schematics illustrate a transmission chain propagating across three different population groups, each depicted by a colored oval shape. Group membership is based on host characteristics or sequence metadata (such as age or geographic region). Each point (circle, triangle or diamond) corresponds to an infected individual, with white filled points indicating sequenced infections, and black filled points indicating infections that are not sequenced. Each diagram illustrates the example of a pair of sequence (white filled points) corresponding to a True Positive, False Positive, False Negative or True Negative. For each of these diagrams, we indicate the corresponding “true” between-group transmission history between the two sequenced individuals and the history inferred from the genomic linkage criterion.

#### Sensitivity

The sensitivity *η*_Δ_ is the true positive rate. It measures how well our linkage criterion captures sequence pairs that reflect the true between-group transmission history. From Table 1, we derive it as:

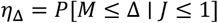

We can compute it as:

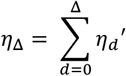

where *η*_*d*_′ is defined as:

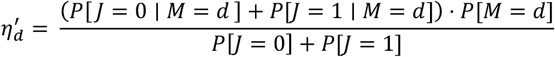

The full derivation is available in Supplementary Information.

#### Specificity

The specificity *χ*_Δ_ is the true negative rate. It measures the fraction of pairs not reflecting the true between-group transmission history that are not captured by the linkage criterion Δ. From Table 1, we can derive it as:

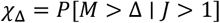

We can compute it as:

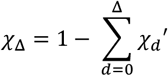

where *χ*_*d*_′ is defined as:

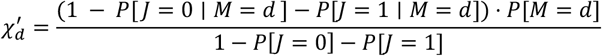

The full derivation is available in Supplementary information.

#### Positive predictive value

The positive predictive value (PPV) *ϕ*_Δ_ is defined as:

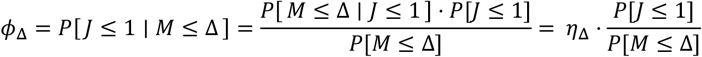

It measures the proportion of linked pairs that correctly capture the between-group transmission history.

#### Accuracy

Similarly, we compute the overall accuracy *A*_Δ_ as:

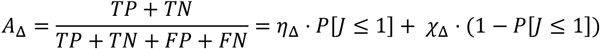

### Spatial and age-based transmission processes’ characteristics

We apply our confusion matrix framework to a combination of pathogens and mixing processes to understand how analyzing the pathogen genome sequences of these different pathogens can provide insights on these population processes. We focus on transmission processes between geographies and age groups. To characterize these mixing processes, we use empirical data to estimate the probability *ω* that a between-group transmission event occurs before a mutation one using mobility and social contact data. This enables us to estimate the probability for a movement of occurring within different geographies in the USA (Table S1) and for a contact of occurring within different age groups (Figure S1-S2) in Washington State. We detail our approach and the data used for this assessment in the Supplement.

#### Relationship between the probability for the infectee to be in the same subgroup as the infector and the between-group transmission event rate λ

We relate the probability *ω* that a transmission event occurs within the same population subgroup to the between-group transmission event rate *λ* using:

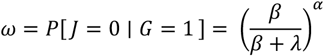

Therefore, for a known value of *ω*, the corresponding between-group transmission rate is equal to:

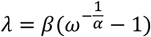

We use *ω* values estimated from mobility and social contact data (see Supplement). *ω* describes the within-group transmission probability per transmission event. For brevity, we refer to it as “within-group transmission probability” in this manuscript.

### Case study across a range of pathogens

#### Evolutionary and transmission characteristics

We apply our confusion matrix approach to Ebola virus, seasonal influenza virus A/H1N1pdm and A/H3N2, measles virus, MERS-CoV, mpox virus, mumps virus, RSV-A, SARS-CoV, SARS-CoV-2 (both pre and post-Omicron) and Zika virus. We assume that sequencing provides whole-genome sequences for all these pathogens. We use previously estimated values of the probability *p* that a transmission event occurs before a mutation one for these pathogens^9^. We explore an additional scenario wherein only the hemagglutinin (HA) segment of the influenza A/H3N2 virus is analysed (corresponding to *p* of 0.92). The shorter genomic target in HA represents a scenario with a reduced per-genome mutation probability.

#### Epidemiological scenarios

We show above that sensitivity, specificity and PPV depend on the distribution of the number of generations separating two individuals picked at random in the population. We use the empirical distributions generated by Wohl et al.^8^ for reproduction numbers *R* of 1.3, 1.5 and 1.8, with results for *R* of 1.3 described in the main text and for *R* of 1.5 and 1.8 presented in Supplementary information (Figures S3-S4).

### Characterizing the parameter space across pathogens

To comprehensively explore trends across pathogens and between-group transmission processes, we apply our confusion matrix approach to a range of evolutionary and mixing parameters. We consider a pathogen with a generation time of mean 4.9 days and standard deviation 4.8 days. This corresponds to the values we used for SARS-CoV-2 (Omicron variant). Assuming a Gamma distributed generation time, this corresponds to a shape of 1.04 and scale of 0.21 days. This parametrization is arbitrary and simply provides a direct way to map evolutionary rates *μ* to values of the probability that transmission occurs before mutation *p* and mixing rates *λ* to values of within-group transmission probability *ω*. We then compute sensitivity, specificity and PPV by varying *p* and *ω* between 0.01 and 0.99 with an increment of 0.01.

### Simulation study to explore the relationship between power and sample size

We evaluate how sample size influences the ability to characterize transmission patterns between groups from sequence data by performing a simulation study using the ReMASTER BEAST2 package^10^.

#### Simulations parametrization

We modelled SEIR epidemics characterized by a basic reproduction number of 2, with a rate out of the exposed (E) compartment of 0.33/day and a rate out of the infected (I) compartment of 0.33/day. This corresponds to a Gamma distributed generation time with shape *α* = 2 and scale *β* = 1/0.33 day. We consider the spread of a pathogen characterized by a probability *p* that transmission occurs before mutation (exploring values between 0.1 and 0.9 with an increment of 0.1) between 4 population groups each of size 50,000. This probability *p* and the generation time’s parametrization as a Gamma distribution translates to a per genome mutation rate of:

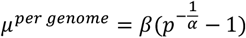

by using similar arguments to those in subsection *Relationship between the probability for the infectee to be in the same subgroup as the infector and the between-group transmission event rate λ*. Assuming a Jukes-Cantor model of evolution, we derive the per site mutation rate (which is used in the simulations) as:

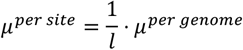

where *l* is the genome length. We run simulations assuming a genome length of 3,000 bp.

We consider transmission processes characterized by within-group transmission probabilities *ω* ranging between 0.1 and 0.9 with an increment of 0.1. We assume a symmetric mixing matrix between groups (detailed parametrization in Table S2).

We assume that a fraction *p*_*seq*_ of all infections are sequenced (exploring values of 0.001, 0.005, 0.01 and 0.05).

#### Relative risk metric performance

To assess the ability of sequences below a given genetic distance threshold to capture mixing patterns, we compute a relative risk (RR) metric which was introduced in prior work and that was shown to capture SARS-CoV-2 transmission patterns between age groups and geographies^2^.

Let *H*_*i,j*_ denote the Hamming distance separating two sequences indexed *i* and *j*, let *S*_*i*_ denote the population subgroup to which sequence *i* belongs. Let *n* denote the number of sequences in the dataset. We define the relative risk 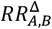 of observing two sequences less than Δ mutations away in population groups *A* and *B* as:

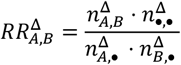

where (using **1** to denote the indicator function):

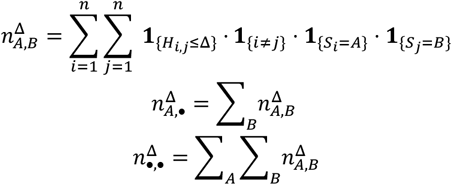

In situations where there aren’t any pairs of sequences observed in either subgroup *A* or subgroup *B* (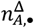 or 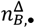 is equal to 0), 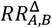 is not defined. To compute RRs even in situations of low pair counts, we rely on a modified RR, defined as:

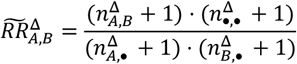

For each combination of *p*_*seq*_, *ω* and *p*, we simulate 50 outbreaks with associated sequence data and compute modified relative risks 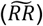 for thresholds Δ ranging between 0 and 15. We then compute the Spearman correlation coefficient between RRs and daily between-group transmission probabilities. In simulations where the modified RRs’ standard deviation is equal to 0, we set the correlation coefficient to 0 (RRs are not informative about between-group transmission probabilities). For each combination of *p*_*seq*_, *ω, p* and Δ, we compute the median correlation across the 50 replicate simulations *ρ*^50^(*p*_*seq*_, *ω, p*, Δ). To characterize the best inference performance for a given sequencing effort *p*_*seq*_, we compute the maximum median correlation across Δ ranging between 0 and 15:

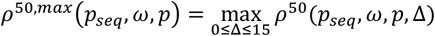

We then characterize the minimum level of sequencing effort required to reach a correlation threshold *τ* (50% and 90%) for each combination of *ω* and *p* as:

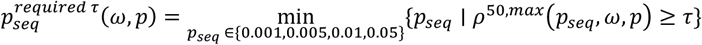

## Results

### Rescaling of evolutionary and mixing rates by generation times

As we rely on genetic distances between consensus sequences, the signal for between-group transmission comes from the occurrence (or lack of occurrence) of mutations between pairs of infected individuals. This signal is determined both by the rate at which mutations accumulate in pathogens genome and the typical delay between successive infections, which defines a window of opportunity for mutations to occur. Because the generation time varies widely between pathogens, the genome-wide mutation rate *μ* thus does not directly map to the expected divergence between transmission pairs (Figure S5A). To account for this, we present our results as a function of the probability *p* that transmission occurs before mutation, which allows to rescale the mutation rate by the generation time distribution and directly captures the expected genetic divergence by transmission pairs (Figure S5B). Figure S5C illustrates how the relationship between *p* and *μ* is modulated by the generation time distribution. We use the same scaling approach to characterize mixing scales by relying on the within-group transmission probability *ω*, which corresponds to the probability that a transmission event occurs before a between-group transmission event. Figure S5D depicts the relationship between *ω* and the between-group transmission rate *λ*.

### Factors impacting the ability of a genetic linkage criterion of capturing transmission between population groups

We find that the linkage criterion’s performance varies across pathogens and is determined by the relative timescale at which mutation and transmission events occur (Figure 3A-C). For example, the sensitivity increases as the probability *p* that transmission occurs before mutation increases (corresponding to slower mutating pathogens, when scaling the mutation rate with the time it takes for each transmission generation to occur), while the specificity and the positive predictive value (PPV) decrease with *p*.

**Figure 3:**
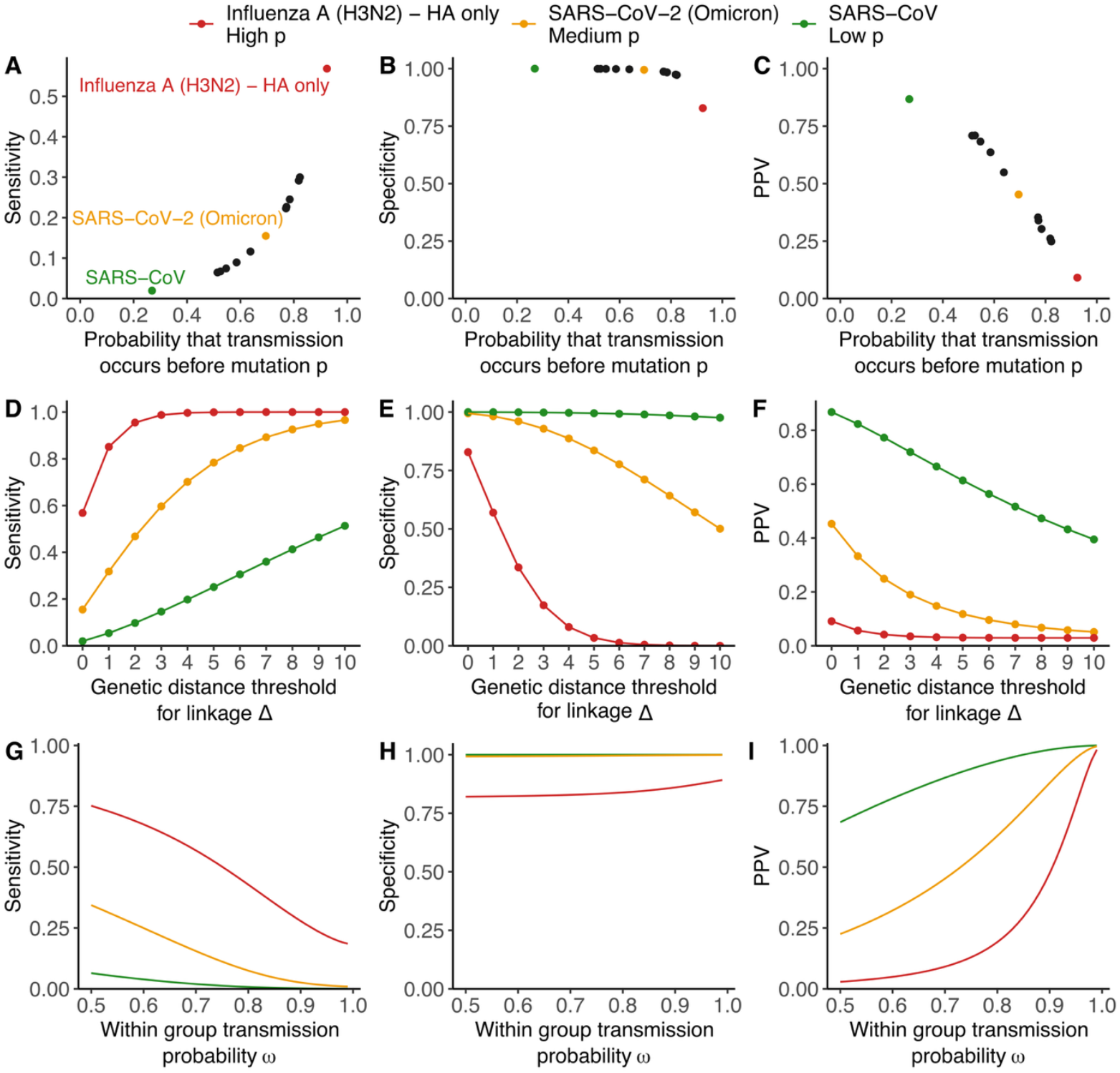
Impact of the mutation and mixing processes and the genetic distance threshold on the sensitivity, specificity and PPV. **A**. Sensitivity, **B**. specificity and **C**. PPV of a linkage criterion defined by Δ = 0 assuming a within-group transmission probability *ω* of 0.7 across pathogens and depicted as a function of the probability that a transmission event occurs before a mutation one across pathogens. **D**. Sensitivity, **E**. specificity and **F**. PPV as a function of the distance threshold used to define the linkage criterion and assuming a within-group transmission probability *ω* of 0.7. **G**. Sensitivity, **H**. specificity and **I**. PPV of a linkage criterion defined by Δ = 0 as a function of the within-group transmission probability *ω*.

To further explore how other parameters impact linkage performance, we focus on a subset of the pathogens depicted in Figure 3A-C. We select this subset to ensure coverage of the full range of *p*: SARS-CoV (low *p*), SARS-CoV-2 (Omicron period – intermediate *p*) and Influenza A/H3N2 (HA only – high *p*). Figure 4D-F depict how varying the genetic distance threshold used to define the criterion impacts overall performance across these pathogens. Specificity and PPV are always maximised at low thresholds while sensitivity increases as the threshold is relaxed. This is expected as a lower threshold captures infections that are more epidemiologically linked and therefore less likely to misrepresent between-group transmission history. These lower thresholds however come at a sensitivity cost, as some relevant pairs aren’t captured by a more conservative criterion.

**Figure 4:**
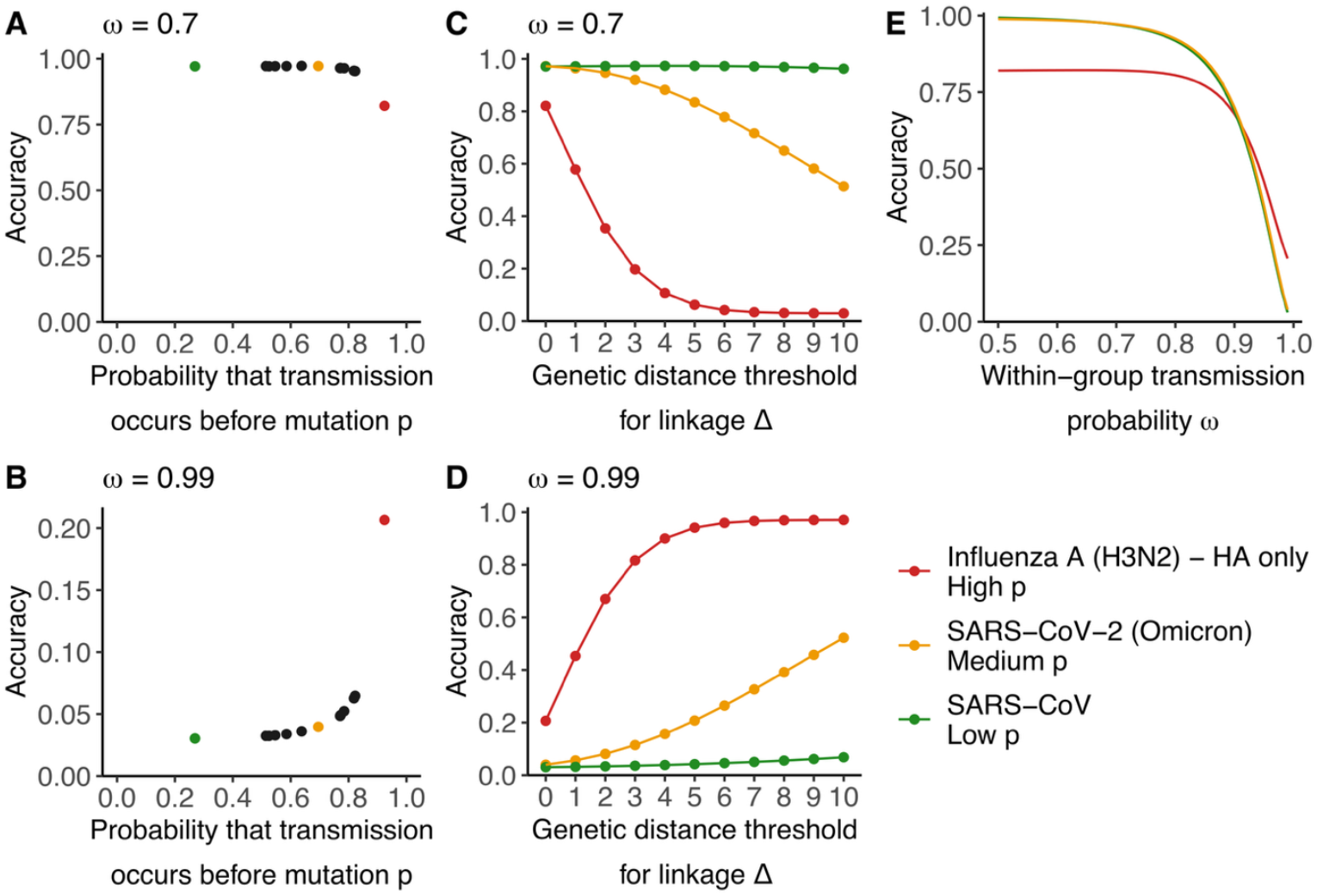
Impact of the mutation and mixing processes, and the genetic distance threshold on the linkage criterion’s accuracy. Impact of the probability that transmission occurs before mutation on the accuracy of a linkage criterion defined by Δ = 0 assuming a within-group transmission probability *ω* of **A**. 0.7 and **B**. 0.99. Impact of the distance threshold Δ on the linkage criterion’s accuracy assuming a within-group transmission probability *ω* of **C**. 0.7 and **D**. 0.99. **E**. Impact of the within-group transmission probability *ω* on the accuracy of a linkage criterion defined by Δ = 0.

We expect the pace at which between-group transmission events occur to impact linkage performance. In Figure 3G-I, we explore the within-group transmission probability’s impact on linkage performance. Faster mixing processes (characterized by lower within-group transmission probability values) are associated with higher sensitivities and lower specificities and PPVs than slower mixing processes, for a specified threshold Δ used to define linkage. This is expected because the probability for a pathogen to have moved several times between groups increases as the within-group transmission probability decreases (faster mixing processes), thereby leading to capturing pairs that are less representative of the between-group transmission history.

We also compute the linkage criterion’s overall accuracy. For lower values of the within-group transmission probability *ω*, the accuracy increases with the probability that transmission occurs before mutation and decreases with the distance threshold Δ, following similar trends as the linkage’s specificity (Figure 4). For higher values of *ω*, corresponding to slower mixing processes, the accuracy follows inverse trends, behaving more similarly to the linkage’s sensitivity. This is expected as the accuracy is computed as a combination of the sensitivity and the specificity, with weights attributed to each metric related to the probability for a pair of sequences of being separated by less than 1 between-group transmission event (see Methods), which depends on *ω*. Unlike PPV that measures the informativeness of linked pairs, accuracy aggregates performance over both linked and unlinked pairs and therefore depends on the frequency of between-group transmission events.

Linkage performance is impacted by assumptions regarding the distribution of the number of generations separating two infected individuals in the population, and therefore the reproduction number. However, a sensitivity analysis varying the reproduction number shows that overall trends are maintained (Figure S4).

Overall, these findings demonstrate that the ability for pathogen genome sequence data to characterize transmission between population groups from genetically proximal sequences depends on the interplay between evolutionary, transmission and mixing processes.

### Limits of genetic sequence data in their ability to characterize population processes

The PPV describes how often a pair of sequences separated by a Hamming distance less than Δ accurately captures between-group transmission history. For a given pathogen (characterized by its probability *p* that transmission occurs before mutation) and transmission process (characterized by its probability *ω* that transmission occurs within the same group), this PPV is highest for a distance threshold Δ of 0 (Figure 3F). To explore the ability of consensus genome sequences in characterizing population processes, we computed the PPV for a threshold Δ of 0 as a function of both *p* and *ω* (Figure 5). To facilitate interpretability, we indicate on the left of the figure how different mixing processes map to within-group transmission probabilities (*ω*) and on the top how different pathogens map to values of *p*.

**Figure 5:**
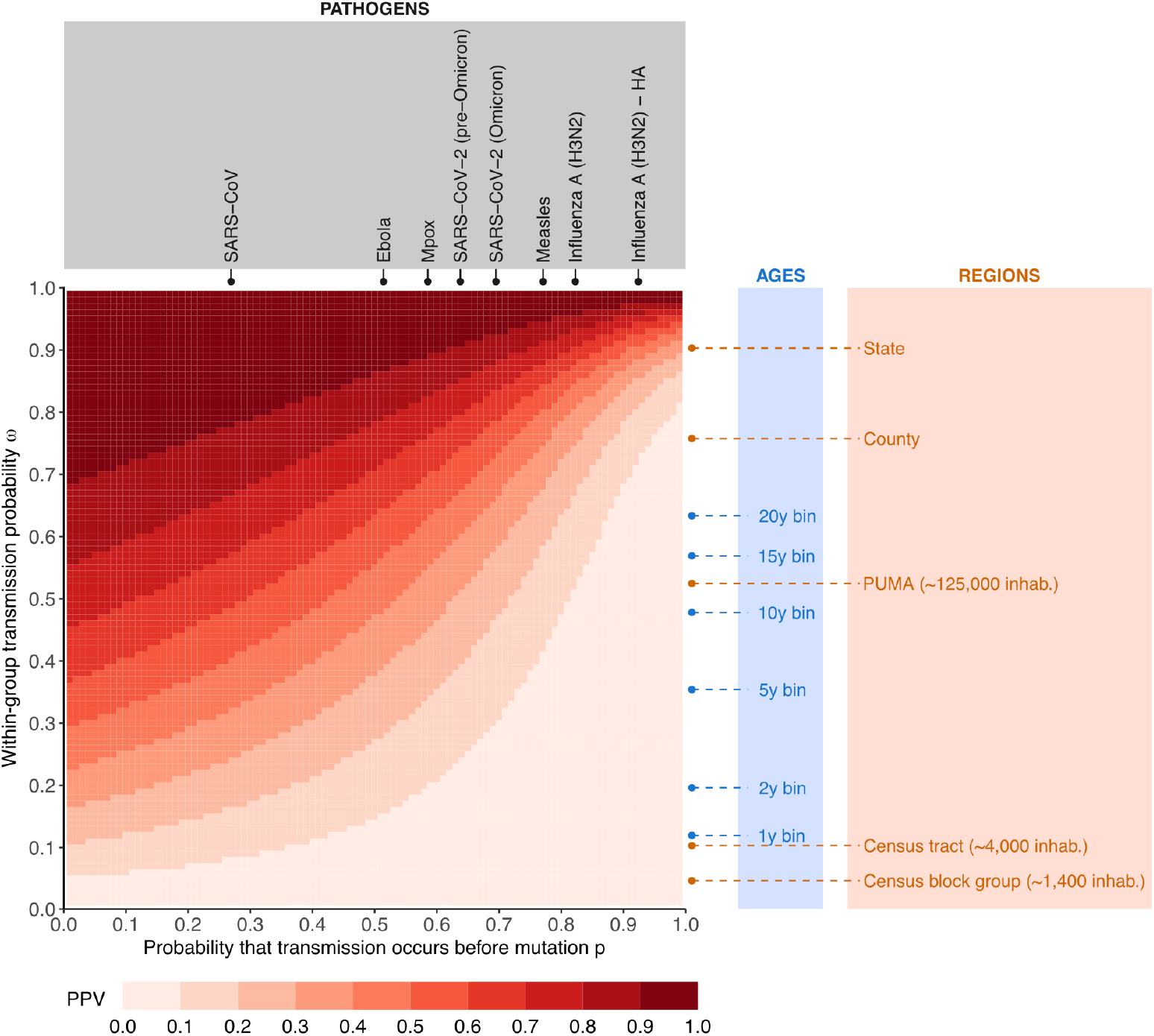
The pace at which mutation and between-group transmission events occur determines the extent to which pathogen sequences are informative about mixing processes. The heatmap depicts the PPV associated with a threshold Δ of 0 as a function of the probability that a transmission event occurs before a mutation one *p* and the within-group transmission probability *ω*. For context, we indicate on the top values for *p* across a range of pathogens and on the right values for *ω* across a range of mixing process. In blue, we indicate within-age group transmission probability *ω* for age groups defined by a range of binning width (1-year binning to 20-year binning) using WA social contact data. In orange, we indicate within-region transmission probabilities estimated using mobile phone movement data for US regions of varying sizes: states, counties, Public Use Microdata Areas (PUMAs), census tracts, and census block groups (CBGs). Details about how we estimate such values are available in the Supplement. Values are computed considering a pathogen with a generation time of mean 4.9 days and standard deviation 4.8 days and a reproduction number of 1.3 (baseline epidemiological scenario).

The PPV for a genetic distance threshold Δ of 0 varies considerably across the parameter space. In our baseline epidemiological scenario, for a pathogen characterized by a *p* of 0.2, we estimate a PPV of 0.28 for a fast-mixing transmission process (*ω* = 0.2) and a PPV of 0.93 for a slower mixing process (*ω* = 0.8). By contrast, these PPVs drop to 0.02 (*ω* = 0.2) and 0.44 (*ω* = 0.8) for a pathogen characterized by a *p* of 0.8. We identify a region of low PPV in the phylogeographic parameter space, primarily in the region wherein values of *p* are higher than values of *ω* (lighter red colours in Figure 5). This corresponds to combinations of pathogens and mixing processes for which analysing consensus sequence data doesn’t provide sufficient resolution to characterize the corresponding mixing process. Each pathogen is therefore associated with a horizon of observability regarding population mixing processes, that depends on the pace at which mutations accumulate within its genome.

Classifying pairs of sequences separated by a genetic distance below Δ (*M* ≤ Δ) but between which no between-group transmission event occurred (*J* = 0) rather as True Negatives rather than True Positives (to focus on between-group transmission events) leads to similar conclusions (Figure S6).

### Trade-off between sample size and positive predictive value

A high PPV ensures the signal from linked sequence pairs is as specific as possible and captures the true between-group transmission history. This PPV is maximized at low distance thresholds Δ, but this comes at a cost of reducing the number of pairs used in the analysis, as lower thresholds result in decreasing linkage probability (Figure 6). This underlines that the performance of our group linkage criterion cannot be considered in isolation from the composition and size of the dataset being studied. A low linkage probability and a high PPV may be preferred in a large dataset but may not be useful when analysing a smaller set of sequences, wherein only a few sequence pairs ultimately meet the linkage criterion. The PPV therefore quantifies the informativeness of genome sequences about population mixing processes in situations where the sample size is large enough for low thresholds not to yield a critically low number of linked pairs.

**Figure 6:**
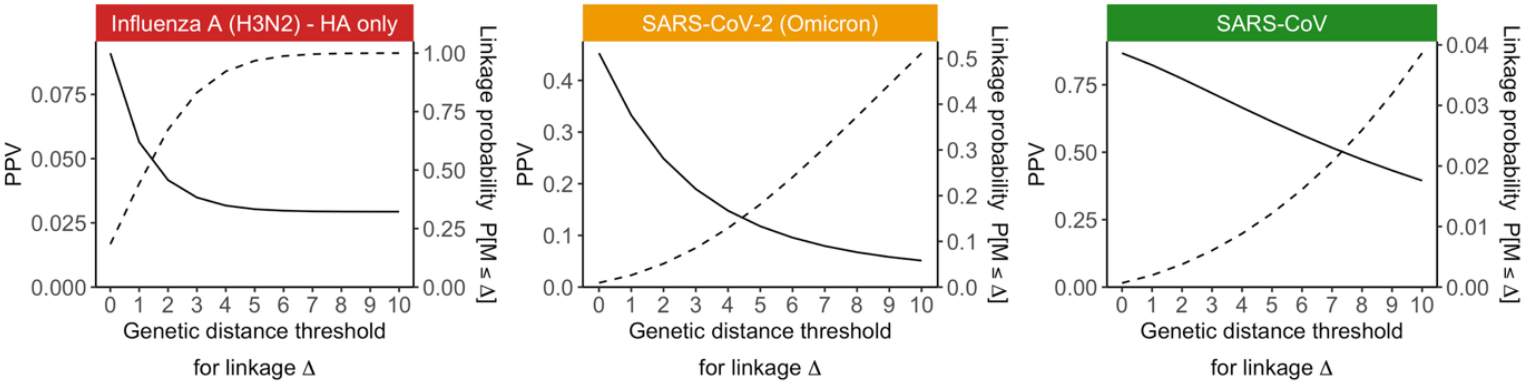
Sample size impacts the distance threshold Δ that maximizes study power. Impact of the genetic distance threshold Δ on the PPV (solid line) and the linkage probability *P*[*M* ≤ Δ] (dashed line) assuming a within-group transmission probability *ω* of 0.7 across pathogens.

To investigate how sample size impacts the ability to characterize mixing processes from pairs of genetically proximal sequences, we simulate synthetic outbreaks and vary the rate at which infected individuals are sequenced. We then compute the correlation between the RR of observing sequence pairs separated by less than Δ mutations and the transmission probability between these subgroups^2^. This RR metric quantifies the extent to which pairs of sequences lying below a distance threshold Δ are enriched in sequences coming from specific subgroups. Prior work demonstrated that this metric could serve as an alternative to traditional tree-based phylogeographic methods^2^ and the median correlation reported in Figure 7 corresponds to a measure of method accuracy.

**Figure 7:**
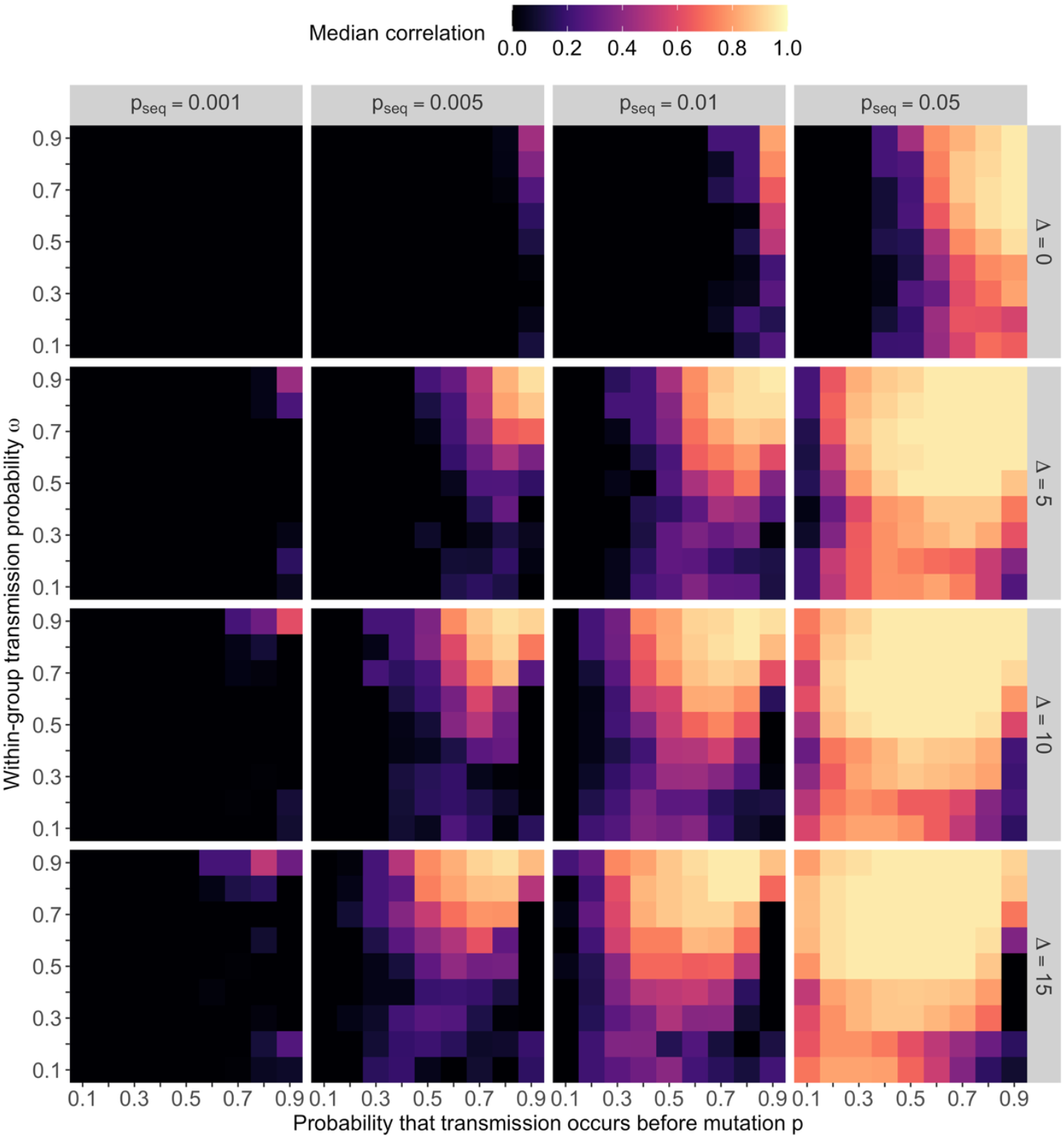
Impact of sample size and genetic distance threshold on inference accuracy. Median Spearman correlation coefficient (across 50 replicate simulations) between the RR of observing sequences less than Δ mutations away (rows) and transmission probabilities between groups, across different sequencing fractions *p*_*seq*_ (columns). Results are displayed as a function of the probability *p* that transmission occurs before mutation and within-group transmission probability *ω*. When the median correlation is lower than 0, we display it as black (corresponding to 0) to improve visualization of positive median correlation values.

Sample size is another important factor influencing study power, with low sequencing fractions being associated with a lower accuracy (Figure 7). Despite the PPV being highest for a genetic distance threshold Δ = 0, relying on this threshold is not sufficient to characterize mixing processes at low sequencing rates (top left facet in Figure 7). Considering less restrictive distance thresholds can increase study power (bottom left facet in Figure 7), by increasing the number of sequence pairs analysed (Figure S7). However, the number of sequences available and analysed imposes an upper bound on inference accuracy, regardless of the distance thresholds (Figure 8A). Figure 5 highlights a fundamental limit for characterizing between-group transmission processes, determined by the relative pace at which mutation and between-group transmission events occurred. Here, we show that study design imposes an additional constraint. While it is theoretically possible to characterize a transmission process characterized by a within-group transmission probability *ω* ∼ 0.5 from a pathogen characterized by *p* ∼ 0.7 (which is similar to assessing SARS-CoV-2 transmission between age groups defined in decade increments) (Figure 5), our simulations highlight that this requires a sufficiently high level of sequencing. For example, in our four-group transmission simulations, sequencing 1% of the infected population would not yield an inference accuracy greater than 90%, and one would need to rely on at least 5% of infections being sequenced to draw such inferences (Figure 8B).

**Figure 8:**
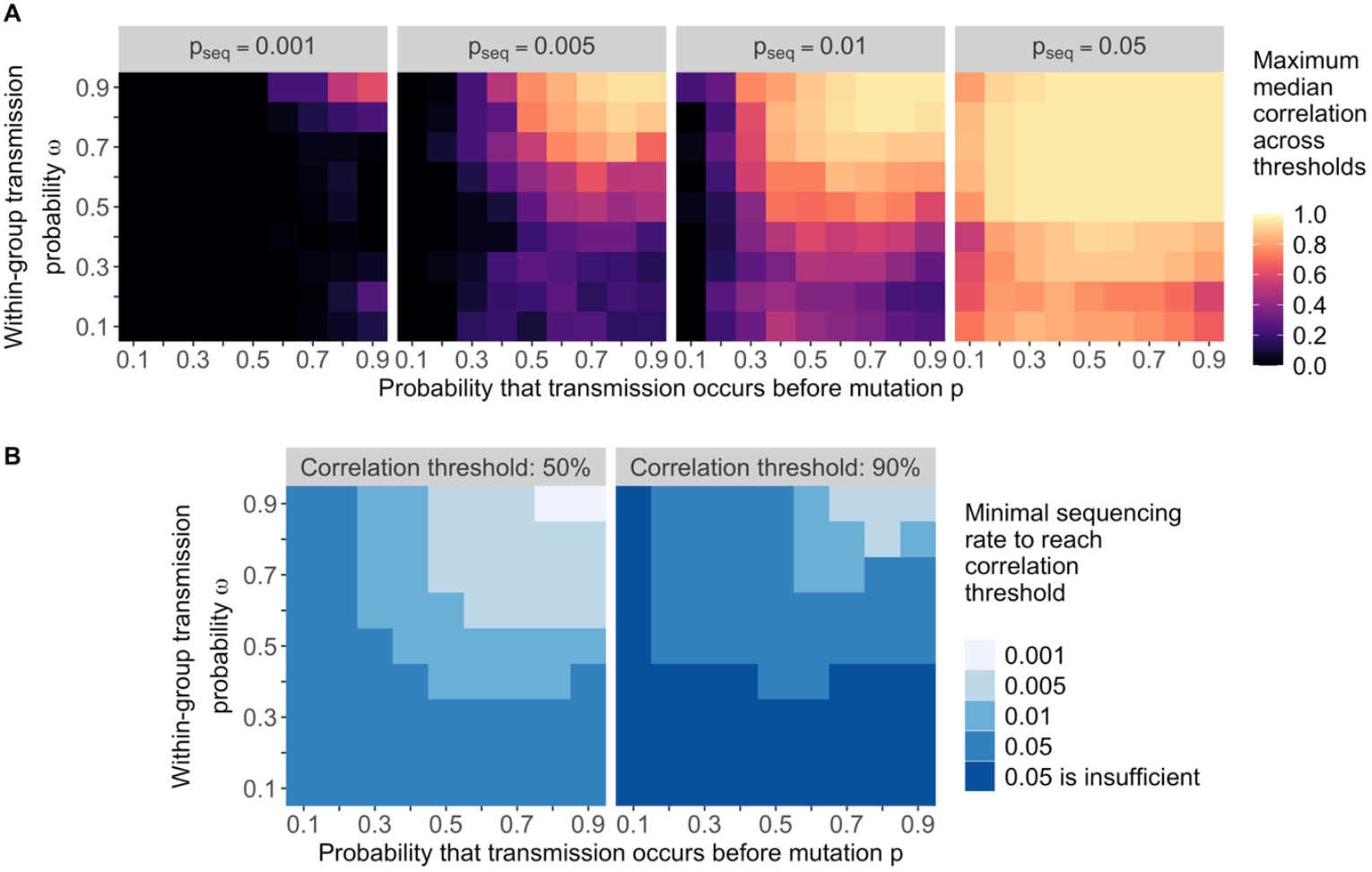
Minimal sampling effort necessary to reach a given inference accuracy. **A**. Maximum median correlation between the the RR of observing sequences less than Δ mutations away and between-group transmission probabilities as a function of of *p* and *ω*, across different sequencing fractions *p*_*seq*_. This maximum correlation is computed across distance thresholds Δ ranging between 0 and 15. **B**. Minimal sequencing fraction *p*_*seq*_ required to reach a maximum median correlation of 50% or 90%, as a function of of *p* and *ω*.

Overall, this shows that the ability to decipher between-group transmission from genetically proximal sequences is influenced by the complex interaction between sampling intensity and the relative timescale at which mutation and between-group transmission events occur.

## Discussion

In this work, we explore how the complex interplay between transmission, population mixing, mutation and sampling impacts the informativeness of sequence datasets about population-level epidemic processes from pairs of proximal sequences. First, we use a confusion matrix approach to quantify whether pairs of sequences are consistent with the underlying between-group transmission history. This succinct formulation enables us to comprehensively explore the space of possible phylogeographic analyses by explicitly incorporating the pace at which mutations accumulate within pathogens genome (measured by the probability *p* that transmission occurs before mutation) and the pace at which pathogens move between groups (measured by the within-group transmission probability *ω*). Our analyses reveal an inherent limit to the resolution genomic data can provide, particularly when between-group transmission events occur faster than the accumulation of mutations within pathogen genomes. Second, we complement this theoretical framework with a simulation exercise to investigate how sampling effort and study design impact the accuracy and power of studies based on clusters of genetically proximal sequences. These simulations reveal a second fundamental constraint, with sparser sampling decreasing the ability of pathogen sequences to decipher transmission between groups.

Our confusion matrix formulation relies on a few assumptions. First, we parametrized the transmission process with a single parameter (the within-group transmission probability *ω*), which we estimate for a few transmission processes (Table S1, Figure S2). In practice the within-group transmission probability varies across groups (Figure S1), socio-demographic settings^11,12^ or with changing immunity profiles. Second, we use the probability that transmission occurs before mutation to describe the probability for an infector and an infectee to have the same consensus sequence. This is a valid assumption for pathogens causing acute infections characterized by narrow transmission bottlenecks^9^. Pairs of consensus sequences from pathogens causing chronic infections also contain valuable information about epidemiological process^13,14^. Determining how the trade-offs we identified translate to such pathogens would be interesting. Finally, we explore how sampling impacts the informativeness of pathogen genome datasets by simulating the spread and sequencing of a pathogen between 4 groups of a population. While we expect the patterns we describe in Figures 7 and 8 to hold for other processes, we couldn’t derive a formula for the sequencing fraction or sample size required as a function of simply *p* and *ω*. Such a quantity would be impacted by factors that are not fully captured by our simple parametrization (including the number of groups, size of groups and between-group transmission rates).

We focus on the information contained by pairs of proximal consensus sequences about transmission processes, to infer a between-group transmission matrix. We thus don’t capture richer information contained by genomic data (such as derived sequences, tree branching patterns and sampling dates). Moreover, pathogen genomes can provide information about other targets, such as introduction patterns, transmission direction, effective population sizes or emergence time, which we don’t explore here. However, we expect the key constraints and trends we identified to be relevant for methods leveraging genomic data differently: if between-group transmission events occur much faster than mutations, genomes will contain little information to characterize the between-group transmission process. This provides a necessary (though not sufficient) condition for the inference of between-group transmission rates from sequence data. Overall, while more sophisticated ways of leveraging genomic data may refine inference about transmission processes, some inherent limitations will persist. Further work directly quantifying such trade-offs for other phylogeographic approaches would be particularly interesting. For example, understanding how the analysis of pairs of consensus sequences could further enable characterizing transmission direction would be particularly relevant. Awareness of such constraints during study design and analysis is critical to avoid false confidence in the resulting inferences. This work emphasizes that the timescale at which between-group transmission occurs imposes an upper bound on the timescale at which genetic variation should be observed to be informative and complements existing literature on the ability to characterize epidemiological processes^3,5–7,15^.

By summarizing between-group transmission with a single parameter (the within-group transmission probability *ω*), we don’t capture the full structure of group mixing. For example, increasing the number of groups at fixed *ω* typically decreases between-group transmission probabilities and increases sample size requirements to fully characterise between-group transmission. Study-specific simulation exercises (similar to the one reported in Figures 7-8) can help define expectations for how binning choices, distance thresholds Δ and sequencing density impact one’s ability to draw inferences. While future work should aim at providing robust guidance on power and sample size requirements to characterize population transmission processes, our conceptual framework provides intuition and identifies actionable levers for modulating the power of genomic epidemiological studies. Sample size and sequencing density in general are major determinants of the power of pathogen genomic studies^8,16^ aimed at quantifying between-group transmission from proximal sequences (Figure 7-8). However, genomic datasets used to perform such analyses are often repurposed from surveillance efforts or studies not initially aimed at quantifying transmission between groups. Increasing sample size to a desirable level might thus be feasible, particularly for retrospective studies. One alternative is to modify the value of key parameters (within-group transmission probability *ω* and probability that transmission occurs before mutation *p*) through study design choices. For example, aggregating individuals into broader population groups both increases *ω* (Figure 5) and decreases the number of between-group mixing rates to estimate. Using WA contact data, we find that analysing age groups in 10-year age bins instead of 5-year ones increases the within-group transmission probability *ω* from 0.35 to 0.48 (Figure 5, Figure S2). Considering spatial spread, aggregating individuals at the PUMA level (around 125,000 inhabitants per PUMA) instead of at the census block group level (around 1,400 inhabitants) increases *ω* from 0.05 to 0.52 (Figure 5, Table S1). The temporal resolution contained in pathogen sequences is also impacted by the analysed genome’s length, as emphasized by prior work characterizing the value of whole-genome trees relative to gene-specific trees in resolving outbreaks in space and time^7^. In our framework, this would be similar to considering a pathogen characterized by a lower *p*. For example, influenza A/H3N2 is characterized by a *p* of 0.82 when concatenating all segments^9^ whereas *p* increases to 0.92 when analysing hemagglutinin segments only. This is congruent with one mutation occurring on average every 19 days across the whole genome versus every 48 days for hemagglutinin segments only.

The inherent limit we identified in analyzing consensus sequences to characterize transmission between population groups underscores the value of developing methods leveraging identical or nearly identical sequences, particularly in settings characterized by rapid between-group mixing. Such methods can enable us to get as close as possible to that limit, and prior work has shown promising results to characterize spatial and social mixing from identical SARS-CoV-2 pairs^2^. However, we showed that, even identical sequences may carry insufficient information to reliably estimate population transmission patterns (Figure 5), particularly when mixing occurs rapidly with respect to mutations. Approaches explicitly leveraging within-host diversity and deep-sequencing (thus capturing faster-occurring evolutionary events) could effectively decrease the value of *p* and have the potential to overcome this limitation.

Overall, our work reveals inherent *horizons of observability* associated with the inference of between-group transmission processes from genomic data that depend on the complex interplay between study design and the relative timescale at which mutation and between-group transmission events occur.

## Data Availability

Code to reproduce analyses is publicly available on GitHub at https://github.com/blab/phylogeo-signal.

## Code accessibility

Code to reproduce analyses is available on GitHub at https://github.com/blab/phylogeo-signal.

## Author contributions

CTK, JL and TB conceived the study. CTK and JL developed the methods. CTK conducted the analyses. ACP and CTK analysed WA mobility data. CTK, JL and TB interpreted the results. CTK wrote the first draft. All authors edited and reviewed the initial manuscript.

## Funding

TB is a Howard Hughes Medical Institute Investigator. This work is supported by NIH NIGMS R35 GM119774. JL acknowledged funding from the Gates Foundation (INV-044865). JL was supported for this work by cooperative agreement CDC-RFA-FT-23-0069 from the CDC’s Center for Forecasting and Outbreak Analytics. ACP is supported by Fogarty International Center’s in-house research division (US National Institutes of Health). CTK and JL would like to thank the Isaac Newton Institute for Mathematical Sciences, Cambridge, for support and hospitality during the programme Modelling and inference for pandemic preparedness, where this work was initiated (EPSRC grant EP/Z000580/1).

## Competing interests

None.

## Disclaimer

The findings and conclusions in this report are solely the authors’ responsibility and do not necessarily represent the official position of the US National Institutes of Health, the Centers for Disease Control and Prevention or the US government.

## Supplementary information

### Supplementary methods

All notations are defined in the main text. To facilitate navigation, a summary of these notations is available in the following table

| Parameter | Description |
| --- | --- |
| *Random variables* |  |
| $J$ | Number of between-group transmission events separating two infections |
| $M$ | Number of mutations separating two infections |
| $G$ | Number of generations separating two infections |
| *Parametrization of the random variables' distribution* |  |
| $\pi_g$ | Probability for two individuals of being separated by $g$ generations |
| $\lambda$ | Between-group transmission event rate |
| $\mu$ | Mutation rate |
| $\alpha$ | Shape parameter for the Gamma distributed generation time |
| $\beta$ | Scale parameter for the Gamma distributed generation time |
| $p$ | Probability that a transmission event occurs before a mutation one. |
| $\omega$ | Within-group transmission probability per transmission event |
| *Parameters associated with the confusion matrix* |  |
| $\Delta$ | Genetic distance threshold used to define the linkage criterion |
| $\eta_\Delta$ | Sensitivity of a linkage criterion defined by a $\Delta$ threshold |
| $\chi_\Delta$ | Specificity of a linkage criterion defined by a $\Delta$ threshold |
| $\phi_\Delta$ | Positive predictive value of a linkage criterion defined by a $\Delta$ threshold |

### Probabilistic framework detailed derivation

#### Distribution of the number of between-group transmission events conditional on the number of generations

This is similar to the derivations made in Tran-Kiem et al^2^ to calculate the distribution of the number of mutations conditional on the number of generations. Let *T*^*evo*^ denote the evolutionary time separating the two individuals we are considering. As the number of between-group transmission events follows a Poisson process of rate *λ*, we have:

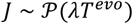

Assuming independence of successive transmission events and because we assumed that the generation time follows a Gamma distribution of parameter (*α, β*), the time between *g* successive generations follows a Gamma distribution of shape *αg* and scale *β*. Let *f*_*x,y*_(·) denote the probability density function of a Gamma distribution of shape *x* and scale *y*. We can derive the distribution of the number of between-group transmission events conditional on the number of generations as:

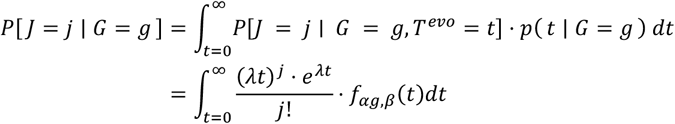

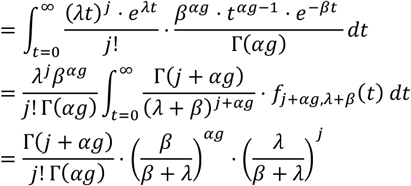

which is the probability mass function of a negative binomial distribution of parameters *r*_*J*|*g*_ = *αg* and 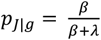.

Therefore:

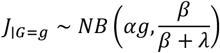

#### Distribution of the number of mutations conditional on the number of generations

By adapting the above demonstration to *M*, that follows a Poisson process of rate *μ*, we have:

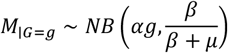

#### Distribution of the number of between-group transmission events conditional on the number of mutations

We introduce *h*(*k*; *r, p*) as the probability mass function, evaluated in *k* of a negative binomial distribution of parameters *r* and *p*. Then, we have:

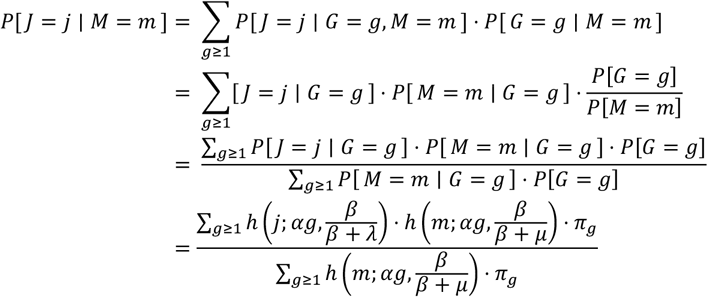

### Confusion matrix parameters derivation

#### Sensitivity

For Δ ≥ 1, we have:

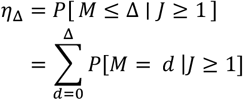

We introduce *η*_Δ_′ as:

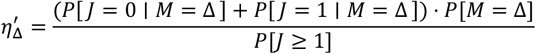

Therefore, for all Δ ≥ 0, we have:

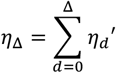

#### Specificity

For Δ ≥ 1, we have:

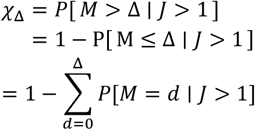

For Δ ≥ 0, we introduce:

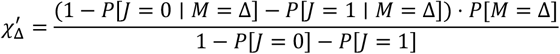

Therefore, for all Δ ≥ 0, we have:

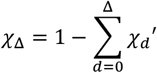

### Characteristics of spatial and age-based transmission processes

#### Age mixing from social contact data

We explore the probability for transmission to occur within the same age group using synthetic social contact data for Washington state from Mistry et al^17^. The latter study provides estimates of the mean daily number of contacts *M*_*i,j*_ that individuals of age *i* have with individuals of age *j* (with one-year age bins). Let *n*_*i*_ denote the number of individuals of age *i*. Age groups can be defined by specifying an aggregation rule. The total number of contacts Γ_*A,B*_ that occur within one day between two population groups *A* and *B* follows:

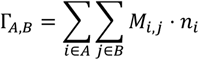

We can also define *c*_*A,B*_ the average daily number of contacts that individuals within age group *A* have with individuals in age group *B* as:

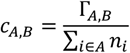

We compute the proportion *p*^*within age*^ of contacts occurring within the same age groups across all contacts occurring within one day as:

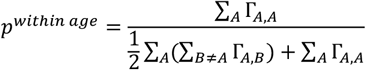

The normalizing factor ½ is used to ensure each contact is only counted once. This metric is a summary statistic of within-group transmission probability at the population level for a specified level of age aggregation. In practice, the probability for a contact of occurring within the same age group is not constant across age groups (Figure S1).

We compute values of *p*^*within age*^ for different binning window size: 1-year, 2-year, 5-year, 10-year and 20-year age bins (Figure S2). For each binning scenario, aggregation starts from age 0 and stops at age 79. We systematically include an age group corresponding to individuals aged 80 and older.

#### Spatial mixing from mobility data

We estimate the probability for transmission of occurring within the same geographical region using mobile device location data from SafeGraph (https://safegraph.com/), a data company that aggregated anonymized location data from 40 million devices, or approximately 10% of the US population, to over 6 million physical places (points of interest, POIs). We use the probability for a movement of occurring within the same geographical unit, while exploring different scales in the US: states, counties, Public Use Microdata Areas (PUMAs), census tracts, and census block groups (CBGs).

At the state and county level, we use data processed in Pullano et al.^18^ and made publicly available in the associated GitHub repository^19^. The authors report the proportion *p*_*i,j*_ of movements of people living in county *i* that travel to county *j*, across different states and for a range of time windows. We focus here on data from January 2020. To estimate the proportion of movements that occur within the same county, we compute a population-weighted average of the proportion of movements occurring within the same county as follows:

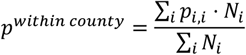

where *N*_*i*_ is the population size of county *i*, derived from US Census data^20^. Using a similar definition, we estimate the proportion *p*^*within state*^ of movements that occur within the same state.

For smaller geographical units (PUMAs, census tracts and CBGs), we rely on a Washington state (WA) focused dataset^21^. We use SafeGraph’s Weekly Patterns dataset to estimate movements within and between WA geographies between January 2019 and June 2022. This dataset provides weekly counts of the total number of unique devices visiting a POI from a particular home location. We restrict our analysis to POIs that are consistently recorded in SafeGraph’s panel throughout the study period.

To measure movement within and between CBGs, we extract the home CBG of devices visiting POIs and limited the dataset to devices with home locations in the CBG of a given POI (within-CBG movement) or with home locations in CBGs outside of a given POI’s CBG (between-CBG movement). This methodology was also applied to census tracts and PUMAs to measure movement within and between these larger geographic units.

To adjust for variation in the size of SafeGraph device panel over time, we multiply raw weekly visits to POIs by a scaling factor, corresponding to the monthly ratio of each CBG, census tract or PUMA’s respective county census population size to the number of devices in SafeGraph’s panel with home locations within that county. We then compute the total number of visits between geographies by summing adjusted weekly counts across POIs, over the entire study period. We use these adjusted counts to compute the proportion of movements occurring within the same county, PUMA, census tract and CBG in WA. Estimated values for the proportion of movements within each geographical unit are detailed in Table S1.

## Supplementary tables

**Table S1:** Estimates of the probability for a movement of occurring within the same geographical unit in the US.

| Geographical unit | Data source | Proportion |
| --- | --- | --- |
| Census block groups | SafeGraph in Washington state | 0.05 |
| Census tracts | SafeGraph in Washington state | 0.10 |
| Public Use Microdata Areas (PUMAs) | SafeGraph in Washington state | 0.52 |
| Counties | SafeGraph in Washington state | 0.81 |
| Counties | Safegraph data from Pullano et al. | 0.76 |
| States | Safegraph data from Pullano et al. | 0.90 |

**Table S2:**
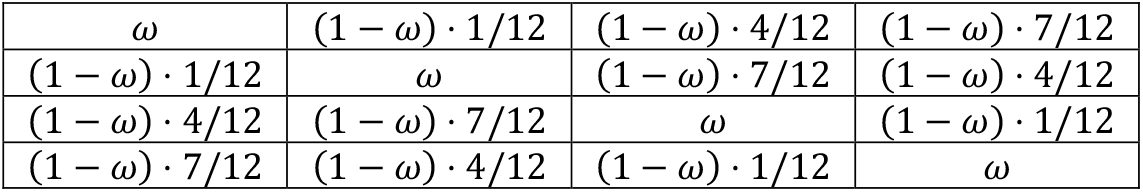
Mixing matrix used in the ReMASTER simulations as a function of the within-group transmission probability parameter *ω* used in the parametrization. Each coefficient corresponds to the probability that an infection coming from someone belonging to group *i* (rows) is in someone in group *j* (columns).

|  |  |  |  |
| --- | --- | --- | --- |
| $\omega$ | $(1 - \omega) \cdot 1/12$ | $(1 - \omega) \cdot 4/12$ | $(1 - \omega) \cdot 7/12$ |
| $(1 - \omega) \cdot 1/12$ | $\omega$ | $(1 - \omega) \cdot 7/12$ | $(1 - \omega) \cdot 4/12$ |
| $(1 - \omega) \cdot 4/12$ | $(1 - \omega) \cdot 7/12$ | $\omega$ | $(1 - \omega) \cdot 1/12$ |
| $(1 - \omega) \cdot 7/12$ | $(1 - \omega) \cdot 4/12$ | $(1 - \omega) \cdot 1/12$ | $\omega$ |

## Supplementary figures

**Figure S1:**
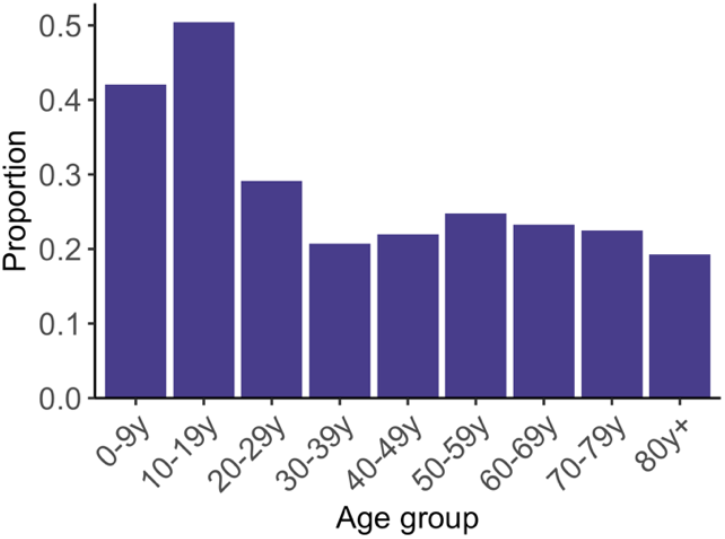
Proportion of contacts occurring within the same age group across age groups, where age group are defined in decades. Estimates were obtained using synthetic social contact data from Washington state^17^.

**Figure S2:**
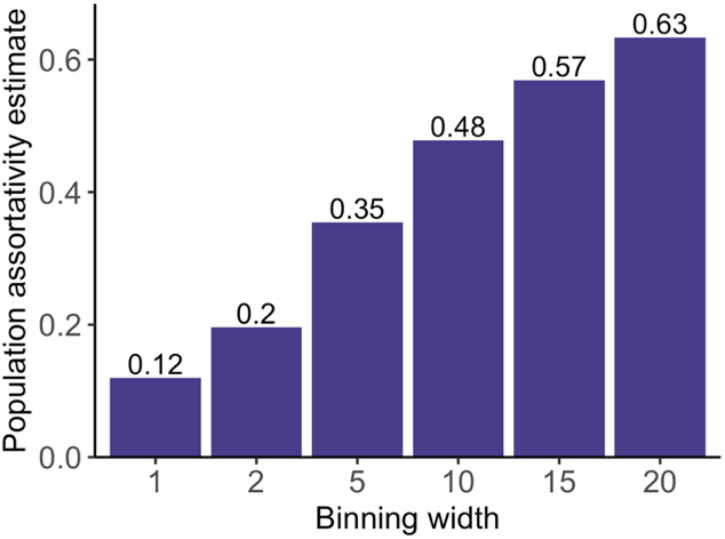
Estimates of the probability for a contact of occurring within the same age group as a function of the binning window width used to define age groups. Estimates were obtained using synthetic social contact data from Washington state^17^.

**Figure S3:**
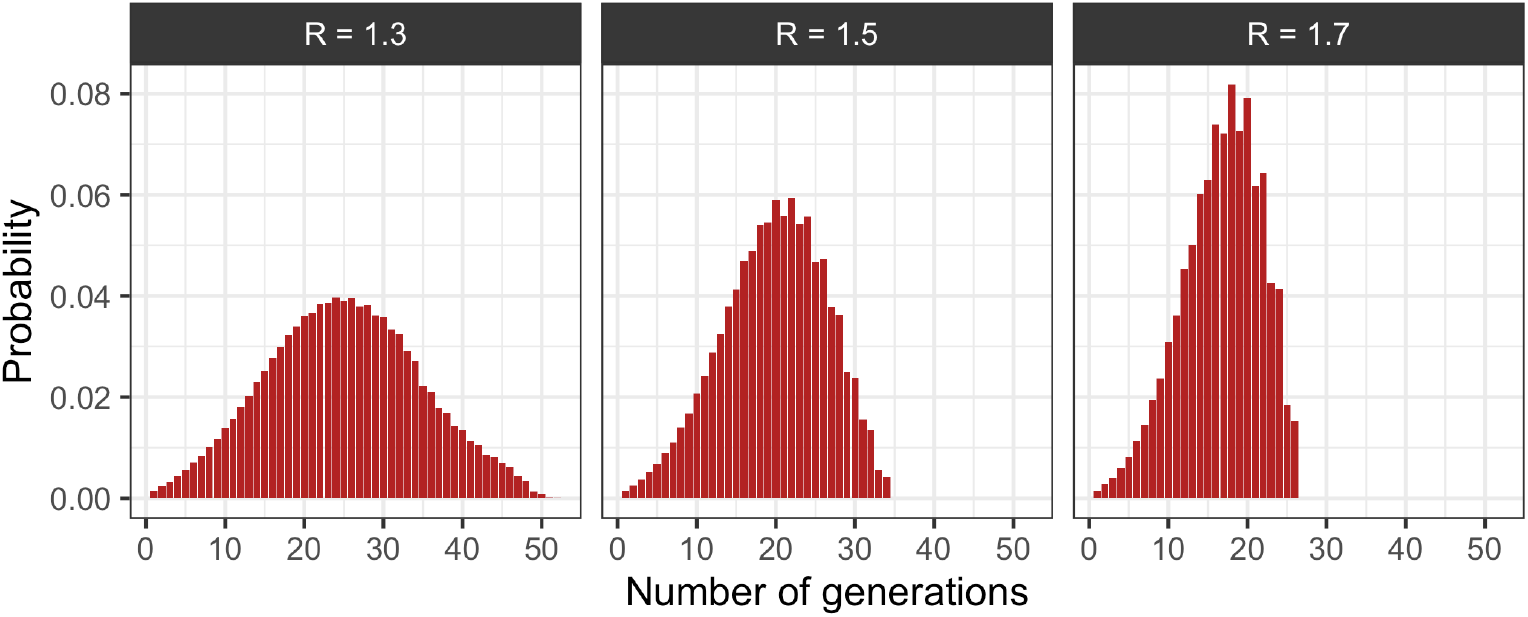
Distribution of the number of generations separating two infected individuals used in the computations. These probabilities are directly extracted from the *phylosamp* R package^22^ as estimated by Wohl et al^8^.

**Figure S4:**
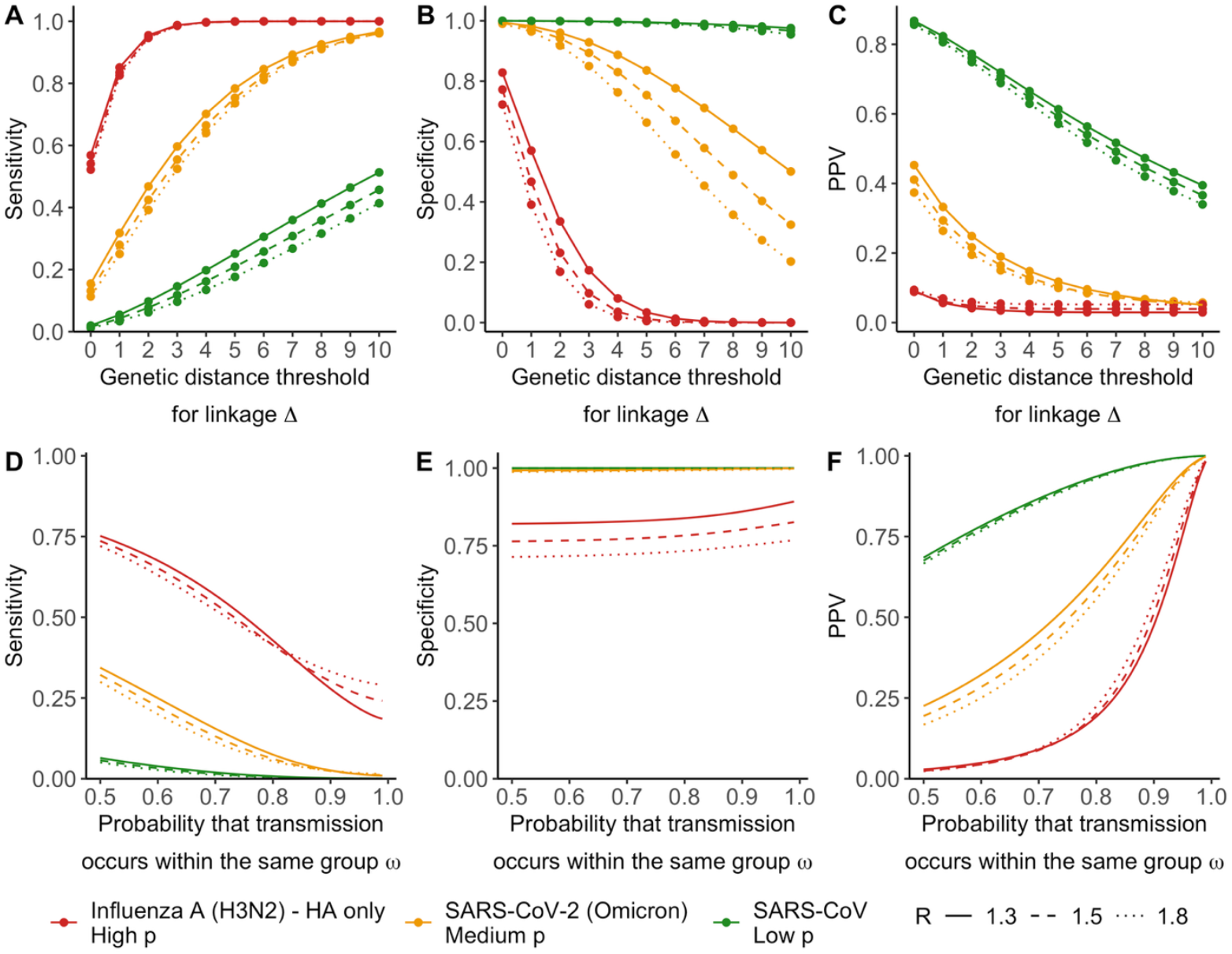
Sensitivity analysis exploring sensitivity, specificity and PPV for different values for the reproduction number R. **A**. Sensitivity, **B**. specificity and **C**. PPV as a function of the genetic distance threshold used to define the linkage criterion and assuming a within-group transmission probability *ω* of 0.7. **D**. Sensitivity, **E**. specificity and **F**. PPV of a linkage criterion defined by Δ = 0 as a function of the within-group transmission probability *ω*.

**Figure S5:**
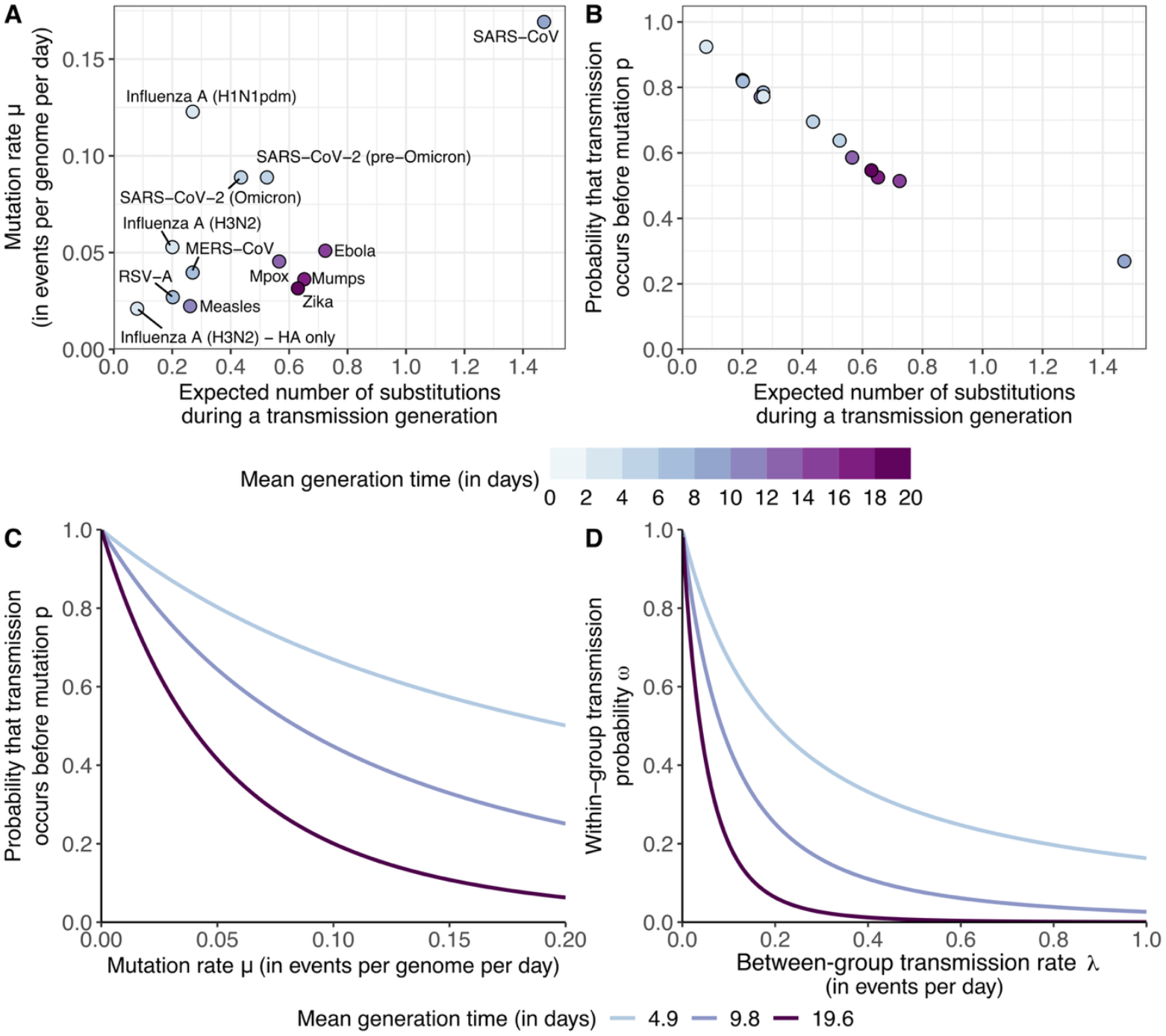
Rescaling pathogen’s evolutionary rates and between-group transmission rates accounting for generation time distribution. **A**. Relationship between the mutation rate *μ* and the expected number of substitutions during a transmission generation across a range of pathogens. **B**. Relationship between the probability that transmission occurs before mutation *p* and the expected number of substitutions during a transmission generation. **C**. Relationship between *μ* and *p* for different generation time distribution parametrizations. **D**. Relationship between the between-group transmission rate *λ* and the within-group transmission probability *ω* for different generation time distribution parametrizations. In C and D, we considered Gamma distributed generation time with the same scale as the one estimated for SARS-CoV-2 (0.21 – see Methods).

**Figure S6:**
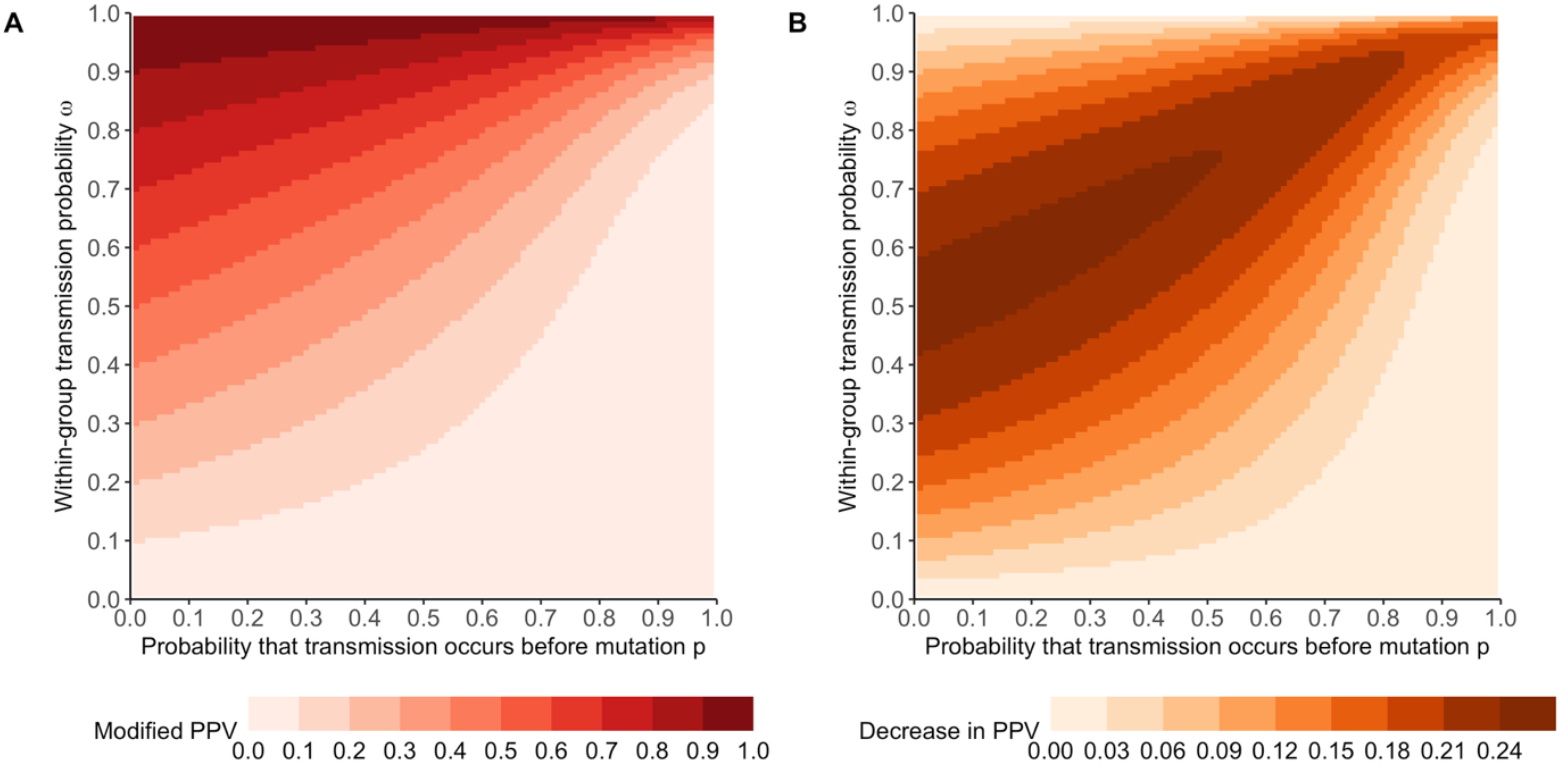
Impact of classifying pairs of sequences characterized by *J* = 0 as True Negatives on the PPV. **A**. Modified PPV as a function of the probability that transmission occurs before mutation *p* and the within-group transmission probability *ω*. **B**. Decrease in the PPV when classifying pairs of sequences characterized by *J* = 0 as TN instead of TP. The modified PPV 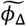 was computed as:

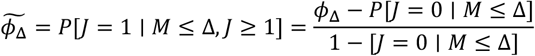

**Figure S7:**
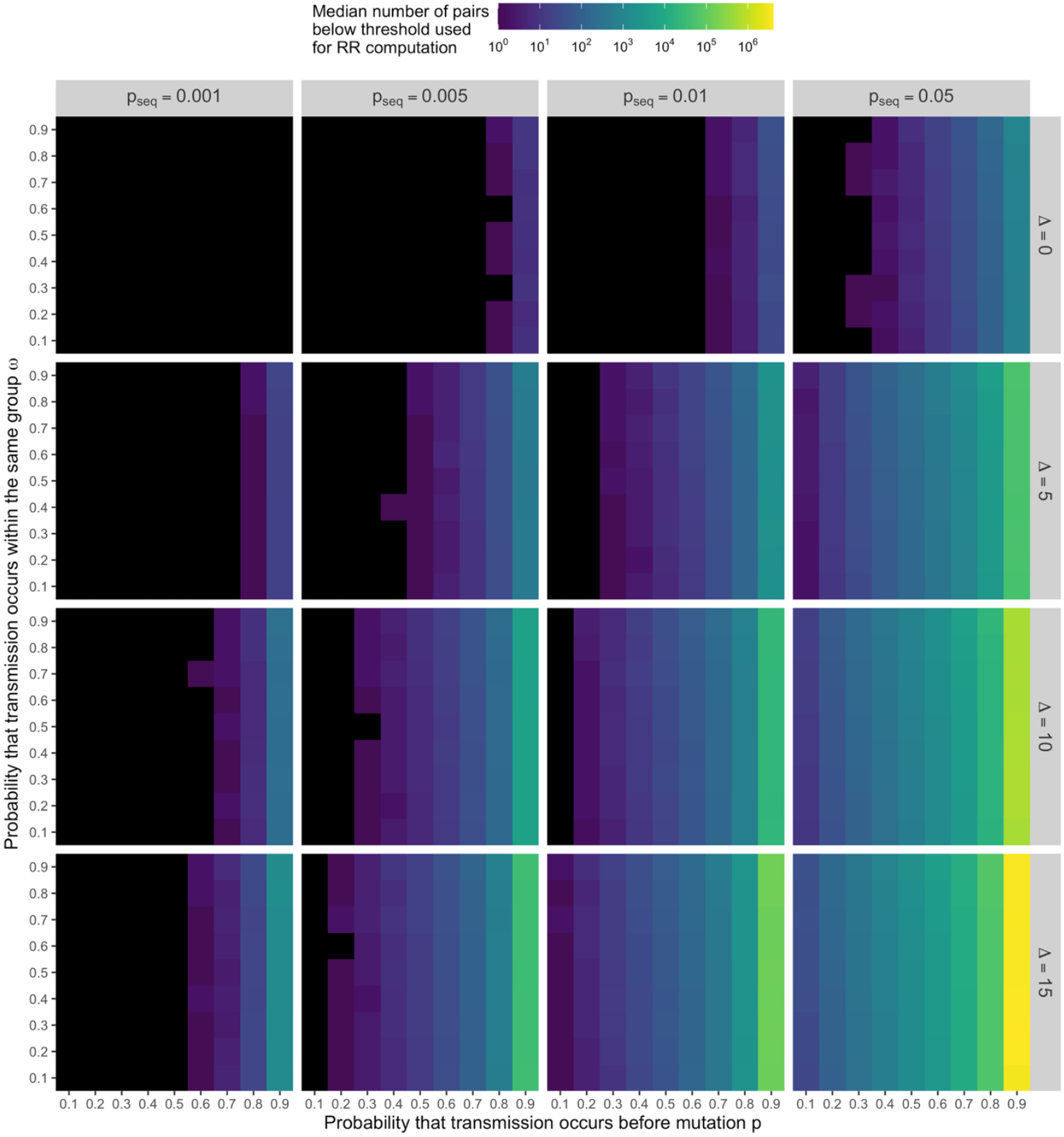
Median number of pairs of sequences less than Δ mutations away across 50 replicate simulations as a function exploring different sequencing fractions *p*_*seq*_ and genetic distance thresholds Δ. Results are displayed as a function of *p* and *ω*. Black tiles correspond to a median value of 0.

## Notes

### Competing Interest Statement

The authors have declared no competing interest.

### Summary of Updates

- Moved former supplementary figure to main (Figure 4) - Rephrasing of the discussion about how applicable our results are in general - Light rephrasing / text editing

